# SARS-CoV-2 detection with de novo designed synthetic riboregulators

**DOI:** 10.1101/2020.07.28.20164004

**Authors:** İlkay Çisil Köksaldi, Recep Erdem Ahan, Sila Köse, Nedim Haciosmanoğlu, Ebru Şahin Kehribar, Murat Alp Güngen, Aykut Özkul, Urartu Özgür Şafak Şeker

## Abstract

Sars-CoV-2 is a human pathogen and is the main cause of COVID-19 disease. COVID-19 is announced as a global pandemic by World Health Organization. COVID-19 is characterized by severe conditions and early diagnosis can make dramatic changes both for personal and public health. In order to increase the reach for low cost equipment which requires a very limited technical knowledge can be beneficial to diagnose the viral infection. Such diagnostic capabilities can have a very critical role to control the transmission of the disease. Here we are reporting a state-of-the-art diagnostic tool developed by using an in vitro synthetic biology approach by employing engineered *de novo* riboregulators. Our design coupled with a home-made point-of-care device setting can detect and report presence of Sars-CoV-2 specific genes. The presence of Sars-CoV-2 related genes triggers translation of sfGFP mRNAs, resulting in green fluorescence output. The approach proposed here has the potential of being a game changer in Sars-COV-2 diagnostics by providing an easy-to-run, low-cost-demanding diagnostic capability.

## Introduction

COVID-19 is a global pandemic caused by a novel coronavirus, SARS-CoV-2, that infects ACE2 positive cells found in the human body. Several tissues and organ systems including the upper respiratory tract, small intestine, liver, and nervous system are susceptible to SARS-CoV-2 infection [1, 2]. Because diverse clinical manifestations of COVID-19 are vaguely defined and share symptoms with other common infectious disease states such as the seasonal flu [3, 4]. Person-to-person transmission and most importantly lack of rapid and adequate testing resulted in loss of opportunity to prevent the spread of the virus in numerous countries at the beginning of the pandemic. As a result, 16,114,449 total cases and 646,641 deaths were reported worldwide as of July 28, 2020 [5].

Test-track-isolate is a fundamental strategy to mitigate the risk overwhelming the healthcare system until a viable therapeutic solution can be produced [6]. Therefore, widespread testing for COVID-19 is vital to block the viral transmission chain. Molecular diagnosis of COVID-19 heavily relied on quantitative real-time PCR (qRT-PCR) that detects the presence of SARS-CoV-2 RNA in nasopharynx, trachea or bronchus, however, testing capacity failed to meet increasing demand as a result of the global shortage in qRT-PCR reagents (i.e. reaction mix, polymerase, reverse transcriptase, etc.) [7, 8]. In addition, qRT-PCR tests generally require a testing facility and trained staff. To compensate for the gap between test number and the pace of infection, many agencies declared an emergency use of *in vitro* diagnostic products for SARS-CoV-2. Consequently, numerous methods, excluding qRT-PCR, were developed for virus detection by targeting its proteins or nucleic acid components [9–12].

Synthetic biology broadens the scope of diagnostic tool alternatives, among which programmable riboregulator toehold switches are straightforward and inexpensive to develop [13]. The versatility of toehold switches enabled researchers to develop biosensors for pathogenic viruses such as Zika [14] and Ebola [15], an antibiotic resistance gene [15] and microorganisms found in the human gut [16]. Combined with the cell-free transcription/translation (TXTL) technology, toehold switches can be used to detect the presence of specific nucleic acid sequences in a point-of-care (POC) device in which the output signal can be a fluorescent protein or the colorimetric degradation product of an enzyme [14]. Furthermore, the limit of detection (LoD) of toehold-based sensors can be increased with a simple isothermal amplification reaction such as NASBA (nucleic acid sequence–based amplification) or RPA (recombinase polymerase amplification) prior to TXTL reaction [17]. Owing to their fast prototyping capability and easy-to-use format, the toehold switch in the TXTL reaction format is an excellent candidate to develop an assay with. Such assay can be implemented in a POC device for surveillance of SARS-CoV-2 in substantial portions of populations, which do not have access to a centralized facility to perform sophisticated diagnosis assays such as qRT-PCR or next-generation sequencing (NGS).

Here, we designed toehold switches to detect the presence of the SARS-CoV-2 virus. Specifically, regions in the SARS-CoV-2 genome, ORF1ab, M, ORF678, and S coding sequences, which are compatible with the NASBA reaction were selected as potential triggers as shown in Figure 1a. Switch sequences specific to the triggers that control expression of sfGFP fluorescent protein were constructed and tested. Two out of four trigger sequences (the regions found in ORF1ab and S proteins) were detected with designed switch sequences both *in vivo* on a genetic circuit and *in vitro* through TXTL reactions. Further characterization assays, i.e. determination of the LoD of switches with clinical samples are required to develop a complete assay, yet we think toehold-based biosensors, implemented in POC devices, will help monitor and control the transmission of the disease, especially in rural and less developed regions.

**Figure 1:**
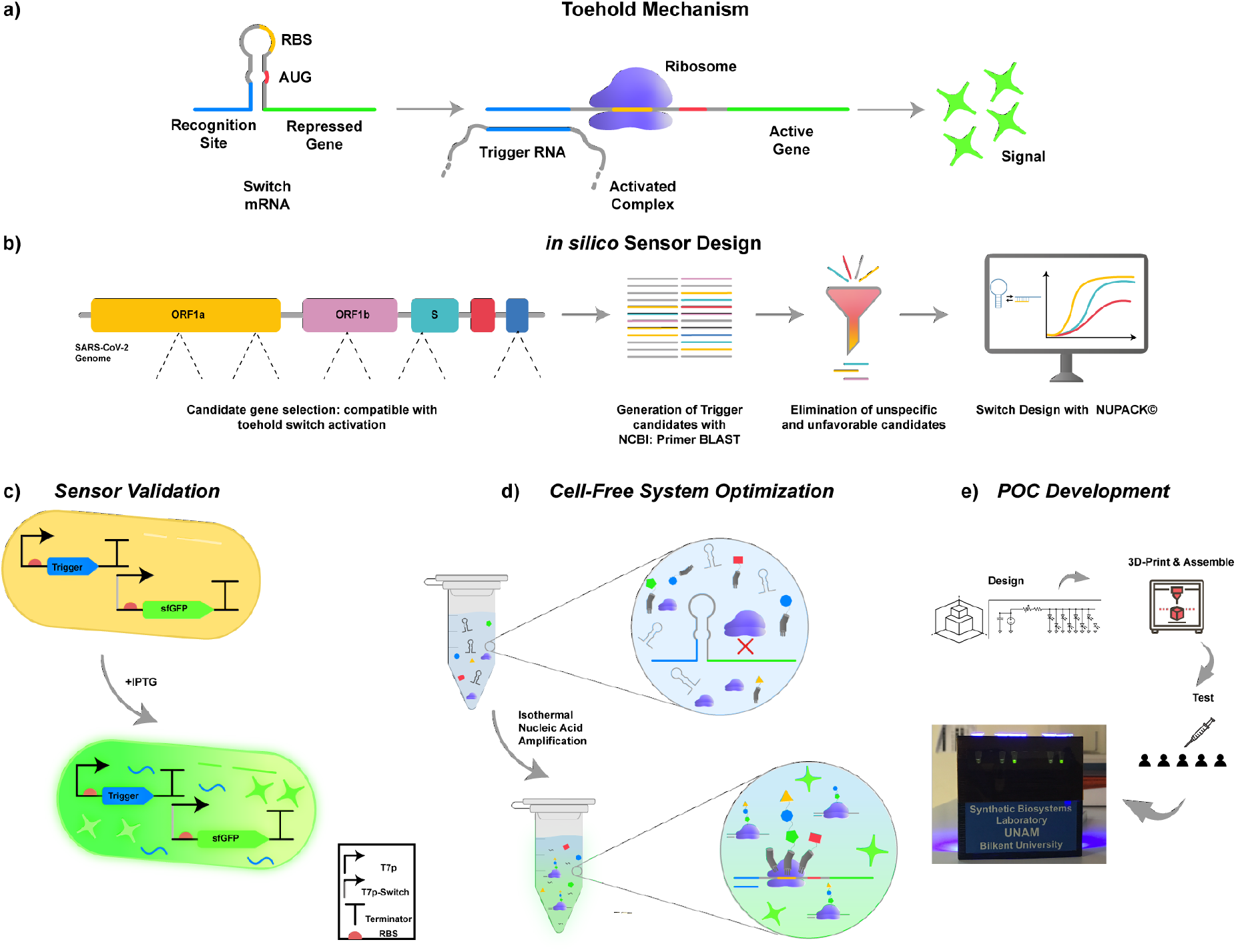
**a)** Schematics of the operating principle of toehold biosensors. **b)** Workflow for cell-free SARS-CoV-2 detection platform. Key target sites for detection of SARS-CoV2 were identified on the viral genome. Toehold triggers are then generated using NCBI Primer BLAST. The trigger candidates that have non-optimal free energy predictions and candidates that are not compatible with species specific amplification were eliminated. Eligible candidates were then used in the design of their respective toehold switches *in silico* using NUPACK©7 software. **c)** Validation of designed toehold switches. Triggers were cloned downstream of T7-LacO promoter. Switches were cloned downstream of T7-LacO promoter and upstream of sfGFP reporter gene. Both constructs were transformed into *E. coli* BL21 (DE3) cells. Cells that only have the switch plasmid and the cells that have both switch and trigger plasmids were induced with IPTG. **d)** Cell-free system optimization. **e)** POC development. Schematics of the development of the portable, low-cost electronic optical reader

## Results and Discussion

We have designed synthetic programmable toehold switch sensors to detect SARS-CoV-2. Toehold switches are synthetic programmable riboregulators that consist of two main components: a trigger and a switch. The switch prevents downstream gene translation through concealment of ribosome binding site (RBS) and start codon through cis-acting RNA interactions. Following the binding of a trans-acting trigger to the switch, RBS and start codon are relieved, resulting in the activation of downstream gene(s) (Figure 1a) [13]. LoD of the toehold switch sensor alone might not be clinically sufficient, hence, primer-based RNA magnification step of trigger region is a requisite to be included before toehold switch sensor detection. Therefore, to screen amplification compatible candidate trigger regions from whole SARS-CoV-2 genome (NC_045512.2), NCBI’s Primer-BLAST tool was utilized. Free energy of secondary structures of candidate triggers were analyzed with Nucleic Acid Package (NUPACK) [18]. Candidates that have free energy over −25.00 (kcal/mol) are selected for further analysis. Selected sequences are aligned using BLAST against the genomes of human and closely related viruses to SARS-CoV-2 to exclusively select SARS-CoV-2 specific triggers. SARS-CoV-2 specific toehold sensors for top five successor triggers are generated using an *in silico* algorithm, based on multi tube design of NUPACK [19] with parameters specified previously [13]. For each trigger candidate, top-ranked switch designs were checked to prevent in frame stop codons. After all of the described filtering processes, overall 18 SARS-CoV-2 specific toehold switch sensors were designed with four trigger sequences (Figure 1b).

To test sensors *in vivo*, fragments for switch and trigger regions were assembled into T7 driven expression p15A and ColE1 origin vectors, respectively. BL21 (DE3) cells were co-transformed with both plasmids and induced with IPTG (Figure 1c). Cells were analysed with a flow cytometer 90 minutes after induction. Furthermore, total fluorescence from cells was measured at 2, 4, 8, and 16 hours following the IPTG addition. Fluorescence measurements indicated that triggers selected from a region in S protein and ORF1ab coding sequence can be utilized to activate switch repressed translation (Figure 2a and 2b).

**Figure 2:**
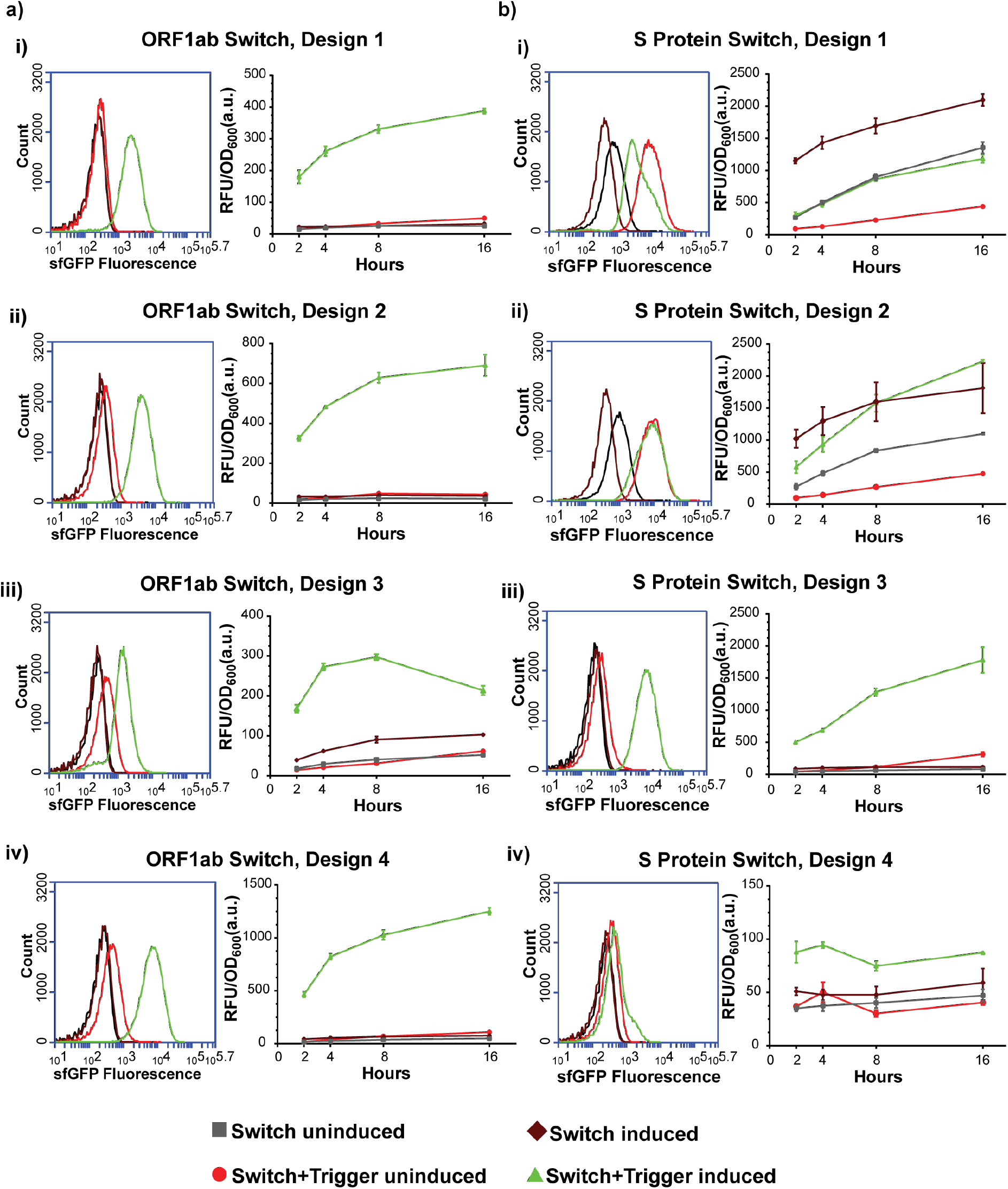
Characterization of best performing two trigger sequences found in S and ORF1ab region *in vivo*. For each trigger sequence, four different unique switch sequences were designed and cloned in p15A vector with T7 promoter and sfGFP coding sequence. Trigger sequences were cloned in ColE1 origin vectors with T7 promoter. Both vectors were transformed in BL21 (DE3) cells and induced. Flow cytometry results were taken after 90 minutes. Meanwhile, cells were monitored for 16 hours via total cell fluorescence measurements using a microplate reader. Cytometer results and total cell fluorescence measurements of designed switch for trigger sequence found in **a)** ORF1ab and **b)** S. For all microplate measurements, three replicates were used.

Based on *in vivo* assessment of all sensors, the top two best performing trigger/switch couples (trigger in S region/design 2 and trigger in ORF1ab region/design 3) were characterized in cell free TXTL reaction (Figure 1d). In reaction set-up, PCR amplified triggers with T7 promoter region and switch plasmids that bear T promoter, corresponding switch sequence and sfGFP coding sequence were used. After addition of DNA molecules, reactions were transferred into a 384-well plate and fluorescence measurement was performed for 6 hours with 10 minutes intervals. Both trigger/switch pairs showed similar kinetics in the TXTL reaction. The reactions were saturated after 180 minutes in terms of total fluorescence. Conversely, ‘switch alone’ circuit did not produce a meaningful fluorescence signal. Activated switch could be distinguished from inactivated and mock controls after 40 minutes incubation (Figure 3a and 3b).

**Figure 3:**
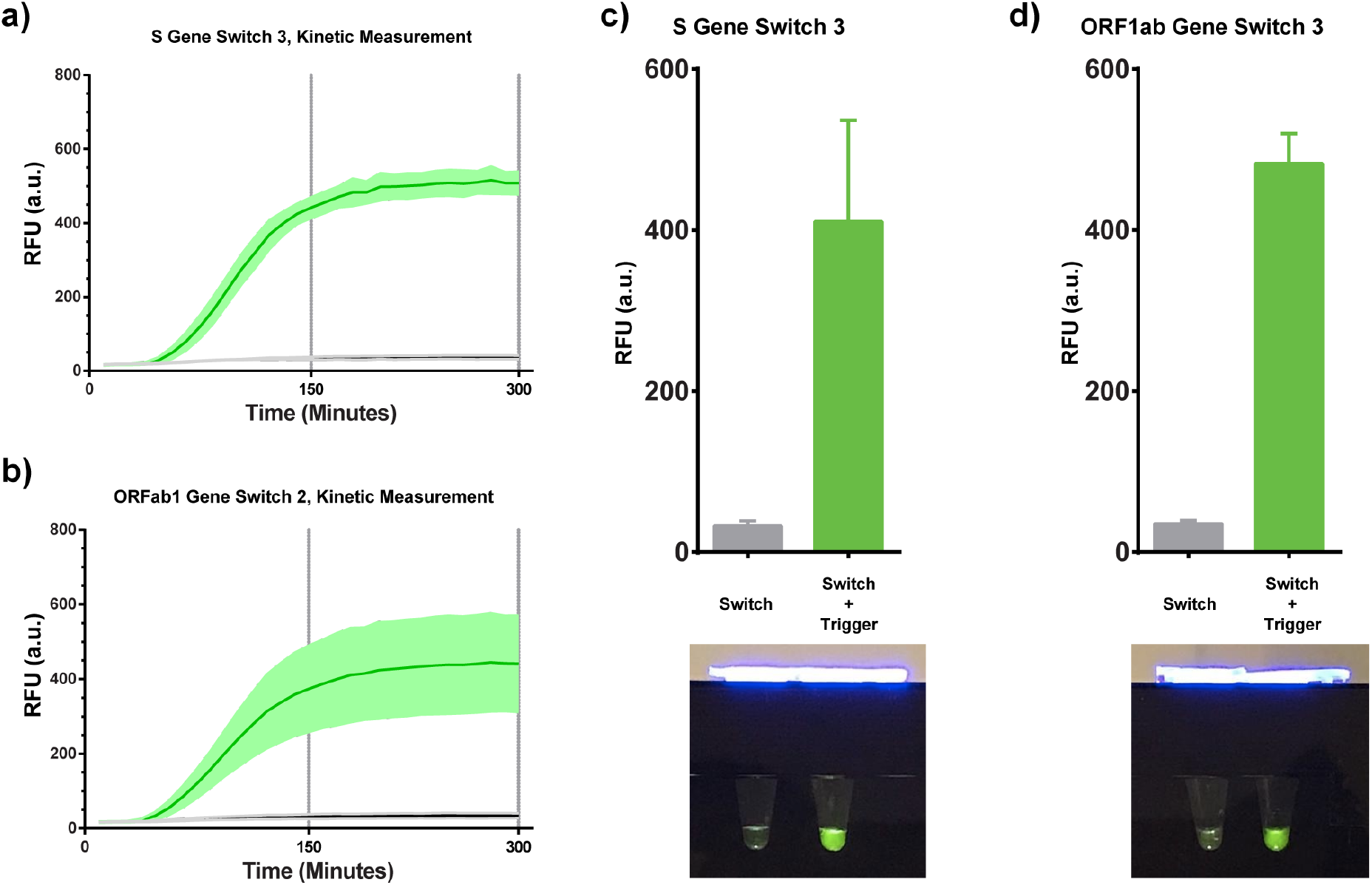
Characterization of S trigger/switch and ORF1ab trigger/switch couple in TXTL reaction. TXTL reactions were set-up in triplicates with addition of PCR amplified triggers and switch vectors. Reactions were monitored for 300 minutes with 10 minute intervals. Fluorescence measurements of **a)** S trigger/switch couple and **b)** ORF1ab trigger/switch couple. **c)** and **d)** Reaction results monitored using a DIY hand illuminator.

A blue light hand illuminator was designed and printed using a 3D printer to visualize the TXTL reaction output. The circuit of the illuminator, based on a modified version of Lucks et al. [20], consists of eight 470nm light emitting diodes (LEDs) connected in series to two resistors: one normal and one variable. The normal resistor is used to limit the current that flows into the LEDs, and the variable resistor is used to adjust their brightness. DIY illuminator powered with three regular 1.5V alkaline batteries was used to inspect the result of TXTL reactions with S trigger/switch and ORF1ab trigger/switch couple (Figure 3c).

## Conclusions

We have successfully developed a *de novo* riboregulator system to detect the presence of the genomic material of Sars-CoV-2 virus. We have employed an *in vitro* synthetic biology tool to develop the diagnostic system. We have screened more than four hundred riboregulator designs and most of them were eliminated due to the poor performance. After designing and testing the system we passed on to the in vitro assays. A region from the spike protein and ORFab1 region of the viral genome were targeted by our designed riboregulators. Our kinetic analysis results showed that the system is capable of detecting the viral genome parts in 40 minutes. Considering the ease of engineering our prosed systems can be re-designed/ engineered to include local mutations.

## Data Availability

We do not have any patient data set.

## Acknowledgements

We thank TÜBİTAK COVID Platform and Bilkent University UNAM for financial support. UOSS thanks to TÜBA, Science Academy, and TÜSEB. We also thank to Can Güven from Bilkent University UNAM for his help with 3D printing of PoC device, and to Zafer Kosar for his help during the initial steps of the study. UOSS also thanks to Ceyda, Ruzgar, Deniz for their invaluable support and understanding during the COVID-19 Pandemic lockdown.

## Materials and Methods

### In silico design

In order to develop an integrated workflow for designing NASBA primers and toehold switches together, a combination of web tools were used throughout the design procedure. For the generation of trigger sequence candidates with primer sites, we used Primer-BLAST (https://www.ncbi.nlm.nih.gov/tools/primer-blast/) by using temperature parameters adjusted to 60ΰC and PCR product size between 120-160 bp. Using the input gene sequences derived from reference SARS-COV-2 genome (GenBank: MN908947.3), we generated a set of trigger candidates with NASBA compatible primer sets for ORFAB1, S, ORF3A, ORFG67, M and E genes including regulatory sequences. Next, we used a web-based analysis tool of NUPACK software (http://www.nupack.org/) to investigate free energies of trigger candidates, and applied a −25 kJ/mol constraint for selecting designs for the next step. Then, by using NCBI-BLAST web tool (https://blast.ncbi.nlm.nih.gov/Blast.cgi), we aligned candidate sequences with six human coronaviruses (229E, NL63, OC43 and HKU1), SARS and MERS to maximize specificity. Candidate triggers that passed, energy and BLAST tests were transferred to the design phase by using the design tool of NUPACK software again to generate switch candidates. 10 candidate switches for each trigger candidate were designed. After completion of the designs, defects, similar sequences and stop codon carrying constructs were eliminated by using a defect function threshold of 25%. Overall, 503 trigger candidates designed with Primer-BLAST, 123 candidates have passed the minimum energy and BLAST requirements, and 5 trigger candidates resulted in up to 4 different switch constructs.

### Assembling Sensor Constructs

For the generation of switch and trigger plasmids, a pET22B backbone (high copy number, AmpR resistance) for triggers; and a pZA backbone (mid copy number, CmR resistance) for switches were used as vectors. In our design, pET22B backbone carries a T7 promoter and a T7 terminator, which can be induced by IPTG addition. On the other side, pZA backbone carries a T7 promoter with sfGFP, superfolder Green Fluorescent Protein gene from the pJT119b plasmid [21] (deposited to the Addgene by Jeffrey Tabor, Addgene #50551) and rrnB_T1 terminator from Registry of Standard Biological Parts at the downstream of sfGFP. Inserts selected during *in silico* design phase were ordered from Genewiz Inc. with Gibson Assembly overlaps. Sequence verification of cloned constructs was done with Sanger sequencing (Genewiz) after the cloning. DH5α strain of *E. coli* was used for all cloning stages.

### Whole-cell Sensor Test

As the first stage of characterizations, switch constructs with a matching trigger sequence were transformed into *E. coli* BL21(DE3) cells with chemical transformation. Meanwhile, standalone switch constructs were transformed as control groups. A single colony from each plate was selected and grown overnight in LB medium at 37ΰC-200 rpm orbital shaker with appropriate antibiotics. Next, cells were diluted in fresh LB medium with antibiotics and induced with 1 mM IPTG at an OD_600_ value between 0.2-0.3. Total fluorescence emission of the trigger plasmid carrying cells and control groups were measured by M5 SpectraMax (Molecular Devices) at 2, 4, 8 and 16 hours.

### Flow Cytometry Analysis

Flow Cytometry measurements were performed by using BD Accuri C6 device (BD Biosciences). Cells were grown and induced as described in the whole-cell sensor test section. For each sample, 300 µL of cells were taken then washed with 1X PBS and diluted as 1/100 before the measurement. Forward scatter (FSC) and side scatter (SSC) thresholds of 10.000 was used to eliminate any background. 100.000 events were recorded for each measurement and the resulting data analysed with BD Accuri C6 Plus software (BD Biosciences).

### Testing Designs with Cell-free Expression systems

Cell-free testing of designed systems was performed by using a commercial cell-free expression kit (NEB, PURExpress) using the protocol provided by the supplier. For each test sample, isolated switch construct plasmid (31.5 nM) and linear trigger DNA (0.34 mM) used. Reactions were measured with M5 SpectraMax (Molecular Devices) for 16 hours to monitor sfGFP expression by tracking fluorescence emission.

### Building a Hand-size Transilluminator

The circuit of the illuminator was designed as mentioned in results part. [20] The illuminator consists of eight 470nm light emitting diodes (LEDs) connected in series to two resistors; one normal and one variable. The case of the illuminator was drawn using Solidworks 2019 then produced with a Makerbot Replicator+ 3D printer using a PLA filament.

